# Epidemiological waves - types, drivers and modulators in the COVID-19 pandemic

**DOI:** 10.1101/2022.01.07.21268513

**Authors:** John Harvey, Bryan Chan, Tarun Srivastava, Alexander E. Zarebski, Paweł Dłotko, Piotr Błaszczyk, Rachel H. Parkinson, Lisa J. White, Ricardo Aguas, Adam Mahdi

## Abstract

**Introduction:** A discussion of ‘waves’ of the COVID-19 epidemic in different countries is a part of the national conversation for many, but there is no hard and fast means of delineating these waves in the available data and their connection to waves in the sense of mathematical epidemiology is only tenuous.

**Methods:** We present an algorithm which processes a general time series to identify substantial, significant and sustained periods of increase in the value of the time series, which could reasonably be described as ‘observed waves’. This provides an objective means of describing observed waves in time series.

**Results:** The output of the algorithm as applied to epidemiological time series related to COVID-19 corresponds to visual intuition and expert opinion. Inspecting the results of individual countries shows how consecutive observed waves can differ greatly with respect to the case fatality ratio. Furthermore, in large countries, a more detailed analysis shows that consecutive observed waves have different geographical ranges. We also show how waves can be modulated by government interventions and find that early implementation of non-pharmaceutical interventions correlates with a reduced number of observed waves and reduced mortality burden in those waves.

**Conclusion:** It is possible to identify observed waves of disease by algorithmic methods and the results can be fruitfully used to analyse the progression of the epidemic.

## 1 Introduction

The COVID-19 pandemic has thrust epidemiology into the public consciousness. Outbreaks, epidemic peaks and waves of transmission are widely discussed. However, we lack a consensus on the definition of many of these terms [31]. Depending upon context, the meaning of the term ‘epidemic wave’ can vary from a well-defined property of a mathematical object through to a loosely defined portion of a time series. Despite definitional problems, using these descriptive terms has value for planning and public health [30]. With Severe Acute Respiratory Syndrome Coronavirus 2 (SARS-CoV-2) believed likely to become endemic [24] and join the ranks of other pathogens which regularly cause epidemics, a reflection on what constitutes an epidemic wave can help to clarify our thoughts and discussion [25].

SARS-CoV-2 has spread across the world after emerging in early December 2019 in Wuhan, China (Figure 1) [21]. Governments across the world implemented non-pharmaceutical interventions (NPIs) with different levels of stringency and speed in an attempt to prevent and/or minimise the importation and local transmission of the virus [26]. Unfortunately, these NPIs often have a substantial associated cost and so it is important to understand how best to reduce transmission in a cost efficient manner. Understanding the epidemic in any single country is a challenge given the myriad potential sources of regional heterogeneities; making meaningful comparisons between countries is even harder. Before we can critically engage in comparative exercises that explore how the size of an epidemic wave relates to factors such as socio-economic status, the stringency of interventions, and other health indicators, we must clearly define what a wave is.

**Figure 1:**
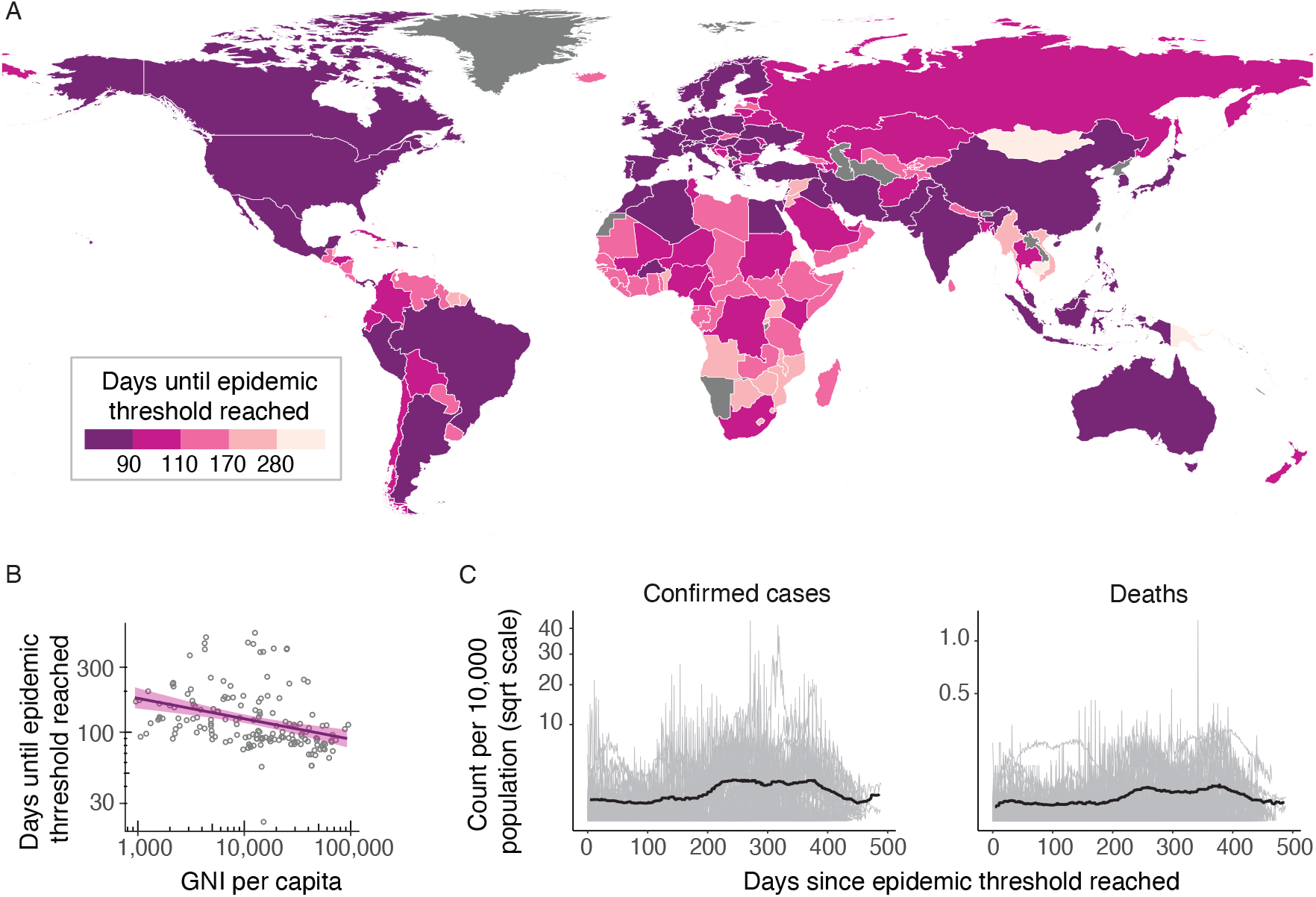
(A) Choropleth showing the number of days since the emergence of the first cases in China on the 31st of December, 2019, until the cumulative number of deaths in each country surpassed 10. Countries with darker colours passed the threshold earlier than the lighter coloured countries. After starting in China, epidemics occurred in Europe, the Middle East and North America before moving south to South America, Africa and the Pacific. (B) Scatter plot showing the correlation between the days until the epidemic threshold was reached in each country against the GNI per capita for that country showing a negative trend, i.e., the pandemic spread to higher GNI per capita countries first. Linear regression line in purple with a shaded 95% confidence interval (C) Time series of the daily number of confirmed cases (left) and deaths (right) per 10,000 population among the countries that have evidence of a second wave (light grey), and the 7-day rolling median of the mean across countries (black line). For each country, the time is taken relative to the date at which the epidemic became established.

In this paper we make three contributions aimed at resolving this issue. First, to facilitate more nuanced discussion, we make explicit the different ways in which researchers use the term ‘epidemic wave’. Second, we provide an algorithm, with a public domain implementation, to partition epidemic time series (of confirmed cases and deaths) into non-overlapping ‘observed waves’, as defined below. We stress that this is not the provision of yet another definition of an epidemic wave; rather it is an exercise in emphasising some of the features any plausible definition should have and making explicit the ones we have chosen. Third, we consider different circumstances that may lead to the formation of observed waves, such as changes in transmission or case ascertainment, and geographic aggregation and in doing so we provide a more nuanced interpretation of the data.

We posit that, when comparing epidemic curves across countries, it is beneficial to do so at the level of observed waves (as identified by our algorithm), in order to remove biases inherent to changing practices over time, such as testing effort and intervention stringency. We provide an example of how to use annotated time series to carry out a global comparative analysis of the influence of NPIs and testing on the size of observed waves.

## 2 Wave detection method

A wave in a time series is informally understood to be a significant, substantial and sustained increase in values followed by a significant and substantial decrease - a temporary increase of at least a certain minimal height which lasts for a certain minimal duration. Both those parameters depend on the context of the problem and, even in a fixed context, practitioners will have legitimate disagreements about whether a given period of a time series is indeed a ‘wave’.

In the context of the epidemiology of infectious diseases, an epidemic wave is a clearly understood feature of mathematical models of transmission dynamics. A wave is characterized by early exponential growth in the number of infected agents, during which the effective reproduction number *Rt >* 1, followed by a saturation phase as *Rt* approaches 1. In an unmitigated epidemic *Rt* = 1 is a threshold condition that corresponds to the moment in which the population reaches herd immunity, after which Rt becomes lower than 1 and cases decrease over time. In systems with mechanisms of fast susceptibility replenishment (loss of immunity) and time dependent transmission modulators (seasonality and epochal evolution), a population can fluctuate around the herd immunity threshold, leading to several epidemic waves over time (e.g. flu epidemics) [11]. In such cases, an epidemic wave can be defined as the period between two local minima. If the model results in the infection rate converging to a stable background level, we can consider the period after the final local minimum to be the final wave of the model.

In the real world, where health surveillance systems are struggling to keep up with an unfolding pandemic, case and death notification definitions are volatile and/or dependent on testing capacity, making it hard to detect when an epidemic wave begins or ends [6]. Furthermore, during the current COVID-19 pandemic, unlike in unmitigated epidemics, changes in social behaviour in response to the development of the pandemic, both as a result of government restrictions and individual choice, have drastically changed the intensity of transmission, Rt, and re-shaped the underlying transmission networks in affected countries. These changes are exogenous to standard epidemiological models, rendering the concept of a wave more ambiguous and raising the need for a clearer discourse around what a wave is, especially given how epidemic waves have been brought to the forefront of public discussion when justifying different levels of policy restriction.

In order to remove the ambiguity surrounding time series of cases and deaths, we propose a robust algorithm for identifying waves (understood as significant, substantial and sustained increases) in time series and apply it to time series of cases and deaths. We do not test for the presence of a significant decrease after the peak, assuming that if a country is currently experiencing an elevated level of transmission it is undergoing a wave which will eventually terminate. As a consequence, this means ongoing waves are still identified.

We describe these as ‘observed waves’, since they differ from the theoretical waves of epidemiological models and, due to measurement error, may also not accurately reflect the underlying SARS-CoV-2 transmission intensity. We consider the essential features of a ‘wave’ and use them to define algorithms which process the many brief rises and falls in case numbers over time and translate them into meaningful ‘observed waves’. For the remainder of the paper, the term ‘waves’ will refer to ‘observed waves’.

Since both cases and deaths can inform the decision about when a wave of COVID-19 begins and ends, we use both series together. On the assumption that the recording of deaths is less subject to change, we take the following approach. If, during a wave in the deaths time series, there is no wave peak present in the cases time series, then we add back a peak of cases which the algorithm had earlier filtered out in order to identify a matching wave of cases. This mitigates concerns about changes in testing over time.

### 2.1 Prominence

Many algorithms exist to identify ‘spikes’ in time series, as this is a key challenge in signal processing [34]. These algorithms identify peaks of brief pulse-like signals. In contrast, epidemic waves are likely to be broad, so that algorithms to identify spikes are not applicable in this context. Instead we aim to identify waves using a prominence-based approach. Prominence can be informally defined, geometrically, as a measurement of how far a peak extends in the vertical direction above the surrounding regions of the time series. The advantage of a prominence-based approach is that it is agnostic as to both the width (i.e., duration) of the peak and the presence of additional local maxima on the shoulder of the peak. This methodology, which stems from recent developments in topological data analysis, has previously been used to analyse medical time series [5, 10].

Prominence may be understood as follows. Consider a fully submerged mountain range, and imagine that the water level is gradually dropping. As the water level falls, ‘islands’ become visible. These are equivalent to portions of the time series that have values above a certain threshold. The global maximum will be seen first, followed by the neighboring values in the time series. As the water level drops further, a second island becomes visible, around the second highest local maximum. As we continue this process, islands will eventually become connected. This occurs as the threshold passes local minima. The ‘prominence’ of a peak is its height above the water level at the point when it merges with an island which originated at a higher peak. (The global maximum could be considered to have infinite prominence, but in this work it is convenient to use the value of the global maximum as the prominence). By considering prominence, we pay less attention to the absolute height of a peak and more attention to its height relative to adjacent peaks. This visual definition is complete, but a definition using mathematical notation is also available in the Supplementary Material.

### 2.2 Algorithm

The wave detection algorithm consists of four parts (A-D, see Fig. [algo]). It processes a time series in order to identify significant, substantial and sustained periods of increased values. These periods are described as waves and the output of the algorithm is a labelling of the peaks of each wave and of the troughs between them.

After initial preprocessing (e.g. smoothing, handling of missing values), a list of all local minima and maxima (critical points) present in the time series is generated. This list corresponds to a set of wave candidates, which are the periods between consecutive local minima. The algorithm then processes this list of minima and maxima, reducing it by removing neighbouring pairs of critical points from the list. This is equivalent to merging two neighbouring wave candidates into one. The conditions on a pair of consecutive critical points to be removed are designed to identify those wave candidates that are insufficiently sustained or substantial and therefore suitable for merging. After the list is reduced, the surviving wave candidates will be of sufficient duration and prominence to be described as waves.

Four sub-algorithms operate to remove pairs from the list of critical points. The operation of each sub-algorithm is described briefly here and illustrated in Figure 2, with more details available in the Supplementary Material and an implementation available at https://github.com/tarunsrivastava145/epidemetrics/. Sub-algorithms A and B ensure that only ‘sustained’ periods of increase are identified as waves. Sub-algorithms C and D ensure that waves are ‘significant and substantial’ by merging waves of insufficient prominence.

**Figure 2:**
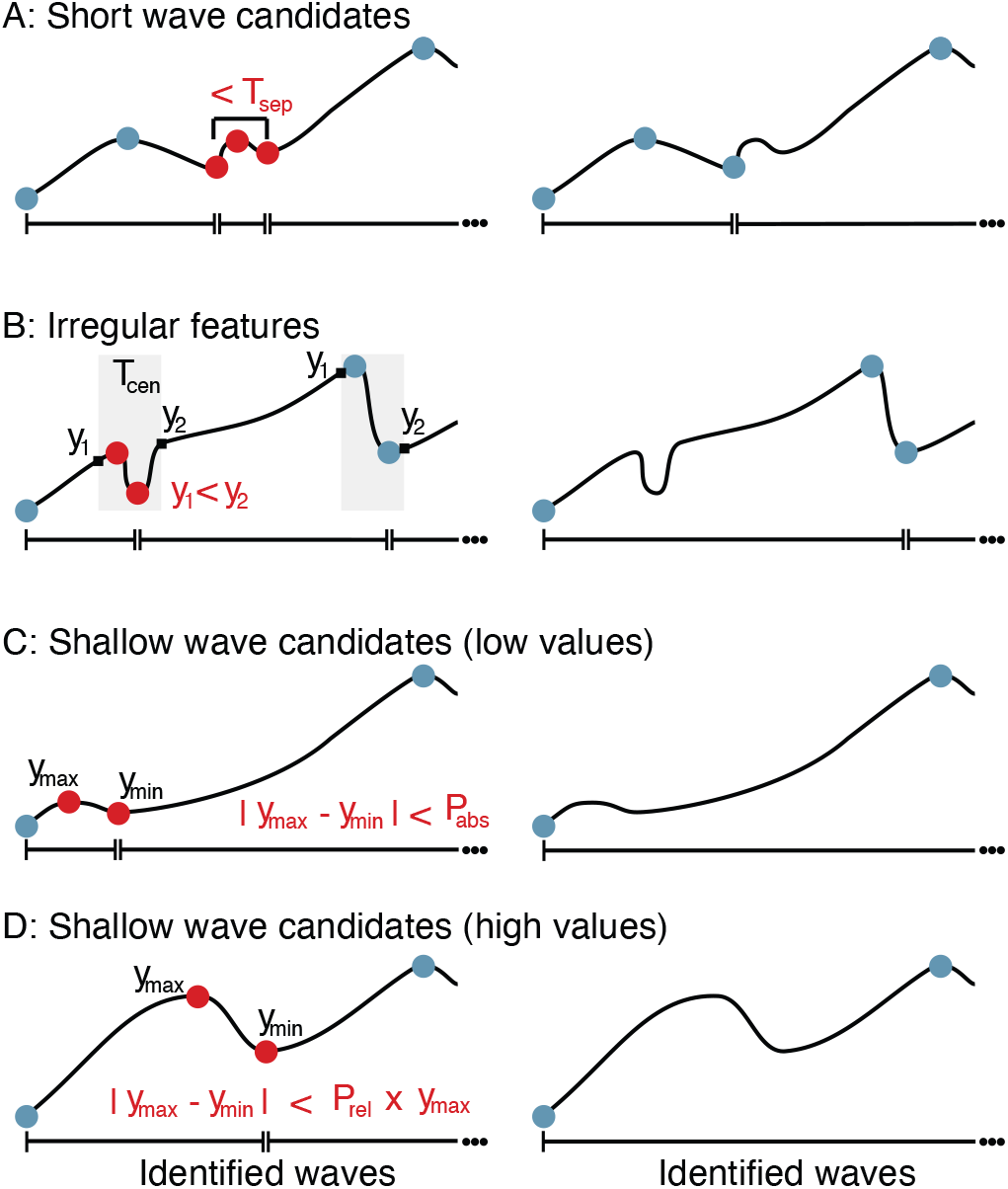
The wave identification algorithm. Time series showing local maxima and minima (solid dots) before (left panels) and after (right panels) filtering with each wave identification algorithm. Red dots represent local minima and maxima that are removed with each algorithm, while those marked with blue dots are retained. Broken horizontal lines beneath each time series show the identified wave durations. Further detail on how each sub-algorithm operates is given in the Supplementary Material.

#### Sub-algorithm A

Merging short wave candidates. For a feature to be described as a wave it must last for a certain minimal period. Wave candidates with duration less than *T*_sep_ are merged into neighbours, in order of increasing prominence. In Fig. [algo] a pair of local minima is separated by less than *T*_sep_. The pair of critical points to be removed is the intervening maximum and the greater of the two neighbouring minima. The result is that the second wave is merged into the third.

#### Sub-algorithm B

Merging irregular features. Where a local minimum and local maximum are separated by a short amount of time, this might indicate either an artifact in the data (for example, it can occur that a large number of historical cases are announced on a single day, creating a spike of cases) or a genuine shift. When two consecutive critical points are separated by less than *T*_cen_*/*2, we centre a time window (shaded) of duration *T*_cen_ on the pair. The data from this period is censored; it is essential that *T*_cen_ *≤ T*_sep_ so that the window contains only a minimum and a maximum. If the feature is transient (as in the first censored window in Fig. [algo], where the feature is a decrease in values but *y*_1_ < *y*_2_), the minimum and maximum are removed. The first wave is merged into the second. However, in the second window the decrease is still visible after censoring (*y*_1_ *> y*_2_) so this pair is retained. The third wave survives this filter.

#### Sub-algorithm C

Merging shallow wave candidates for low values. Waves with prominence less than *P*_abs_ are merged into neighbours. The prominence of the maximum in red (which is *y*_max_ ™ *y*_min_), does not exceed *P*_abs_. The maximum is therefore removed along with the greater of the neighboring minima, so that the first wave is merged into the second. When values are higher, this sub-algorithm is redundant, as sub-algorithm D would also merge all low-prominence waves.

#### Sub-algorithm D

Merging shallow wave candidates for high values. Case numbers may not fall sufficiently after a peak for a wave to be clearly separated from a neighbour. Waves with prominence less than *h · P*_rel_, where h is the value at the peak, are merged into a neighbour. This assumes a background level of 0 for the time series, which may not be suitable for other applications. In the illustration we have set *P*_rel_ = 0.61, which is the value used in this work. The prominence of the maximum in red (which is *y*_max_ − *y*_min_), does not exceed 0.61 *· y*_max_. The maximum is therefore removed along with the greater of the neighboring minima, so that the first wave is merged into the second.

### 2.3 Parameters

Certain parameters must be set by the user in order to determine how substantial or sustained a feature must be before it is identified as a wave. These are *T*_sep_, *T*_cen_, *P*_abs_ and *P*_rel_. The choice of *T*_sep_ can be informed by biological information on the transmission of the disease. For example, epidemics that occur over a longer period of time such as tuberculosis and HIV will require a longer value for *T*_sep_ than more dynamic epidemics such as measles. The motivation for sub-algorithms A and B is to remove local fluctuations which are due to changes in things beyond the transmission process and hence this parameter must be informed by the natural timing of that transmission process. *P*_abs_ and *P*_rel_ should be set large enough to exceed the level of noise in the data and also meet the user’s subjective opinion of what level of cases should be considered substantial and how well separated two waves must be.

The parameters used in this paper are set out in Table 1. Given that the serial interval of COVID-19 is less than five days [23] and the mean incubation period is also approximately five days [21], we find setting *T*_sep_ = 35 d and *T*_cen_ = 35 d to be a reasonable parameter choice. *P*_abs_ and *P*_rel_ were chosen as the output provided a suitable match to expert opinion. *P*_abs_ is set to vary with the population of the country, but with a floor to account for noise and a ceiling for sufficiently large countries.

**Table 1:** Parameters used in the Wave Identification Algorithm. For cases *P*_abs_ = min(max(45, Population *·* 3.3*/*1, 000, 000), 500) and for deaths, *P*_abs_ = min(max(7, Population *·* 1*/*1, 000, 000), 70).

| Parameter | Definition | Value (Cases) | Value (Deaths) |
| --- | --- | --- | --- |
| $T_{\text{sep}}$ | Minimal wave-length, i.e. separation between consecutive minima or maxima. | 35 days | 35 days |
| $T_{\text{cen}}$ | Each minimum and maximum separated by less than $T_{\text{cen}}/2$ are tested in Sub-algorithm B to determine if they persist over a period of duration $T_{\text{cen}}$. It is required that $T_{\text{cen}} \leq T_{\text{sep}}$. | 35 days | 35 days |
| $P_{\text{abs}}$ | Absolute prominence threshold, defined as the minimal prominence for local maxima of waves. | See caption. | See caption. |
| $P_{\text{rel}}$ | Relative prominence threshold, defined as the minimal prominence for local maxima of waves as a proportion of the value at the local maximum. | 0.61 | 0.65 |

The final output is a list of local maxima and minima dividing the total period of the time series into waves. All remaining waves have prominence exceeding *P*_abs_, length at least *T*_sep_, and there is a long (*> T*_sep_) and substantial (*P*_rel_ = 61%) drop between any pair of peaks. The algorithm will indicate that no wave was detected if the time series never exceeds *P*_abs_.

### 2.4 Data and Preprocessing

We extracted data from the OxCOVID19 Database [22], which contains geographically unified information on epidemiology, government response, demographics, mobility and weather at a national and sub-national level collected from various sources. From this database we selected epidemiological data from the European Centre for Disease Prevention and Control [1] and to track measures imposed by governments we used data curated by researchers from the Blavatnik School of Government, University of Oxford [Hal2020]. Information on testing was obtained from the Our World in Data COVID-19 dataset [27]. This covers 103 countries and provides the total tests carried out as well as new tests per day. Note that the statistics provided by each country may differ in whether they refer to the number of tests conducted or the number of individuals tested. Data was last extracted on 23 June 2021, with a cutoff for time series data on or before 31 May 2021 to account for incomplete data due to delays in reporting.

Prior to analysis of the epidemiological data, missing daily values for cumulative confirmed cases and deaths were filled in by linear interpolation. The number of new cases and deaths per day was then computed as the difference between the cumulative values for successive days. Negative values were replaced with the last non-negative observation. Such values commonly arise when reporting authorities subsequently correct their figures for total cases or total deaths. After this initial cleaning substantial stochasticity is still present in the time series, due to factors such as backlogs in the number of cases over weekends and errors in consolidating municipal sources. To better understand the underlying trend and identify significant waves, we computed a fourteen-day moving average to smooth the data.

The data series for the number of performed tests is provided with this seven-day smoothing already performed and we use the smoothed value directly [27]. We computed the positivity rate as a ratio of new cases per day to new tests per day (using raw data in both cases), and subsequently smoothed this rate with a seven-day moving average. Earlier in the time series positivity rates greater than 1 occasionally arise; these were removed. From the Blavatnik School data [13], we isolated the ‘stringency index’, a measure of the overall stringency of government interventions, as well as the dates at which various flags were raised indicating the implementation of restrictions, such as the ‘C3’ flag indicating the mandatory cancellation of public events.

## 3 Results and Discussion

After clarifying the different ways in which the term ‘waves’ is used by different communities, we proposed a novel algorithm for identifying epidemiological waves in time series of cases and deaths. The algorithm delineates significant, substantial and sustained periods of increased case numbers, separated from each other by significant and substantial decreases.

### 3.1 Identification of epidemic waves of COVID-19

The algorithm was applied in the context of COVID-19 for every country for which data was available. By applying the algorithm to both the cases time series and the deaths time series, we could use cross-validation of the results to address the confounding effect of changing case ascertainment and better identify the waves of cases.

Figure 3 gives a sample of how the different layers of the algorithm merge wave candidates together.

**Figure 3:**
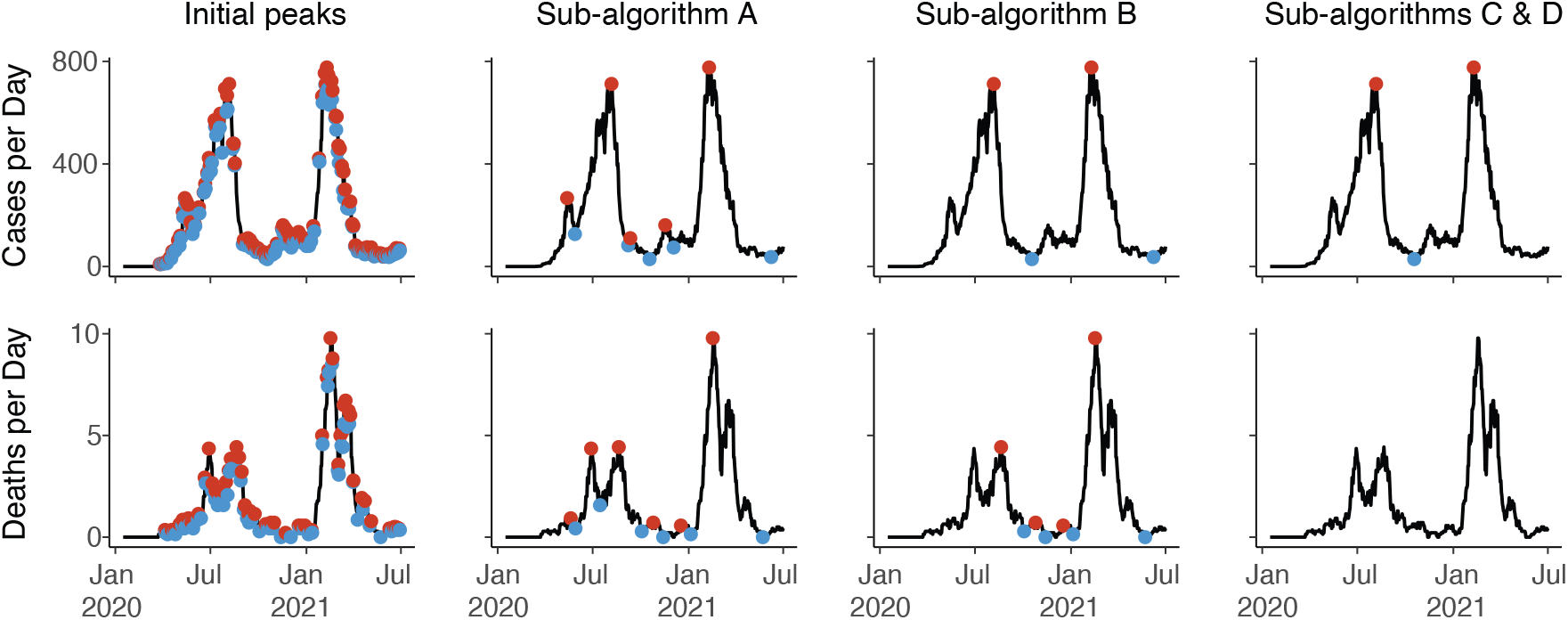
Illustration of how each Sub-Algorithm operates to reduce the list of identified minima (solid blue circles) and maxima (solid red circles) in the time series of cases and deaths for Ghana. In black, the smoothed data for each time series is shown. Solid circles indicate which minima and maxima have survived the pruning process at each stage.

Figure 4 shows some characteristic examples of time series studied along with the division of those series into waves. It is clear how decisions in the setting of parameters control which features are removed as noise and which structures are combined to be considered as one wave. The utility of cross-validating between the cases and deaths time series is also demonstrated.

**Figure 4:**
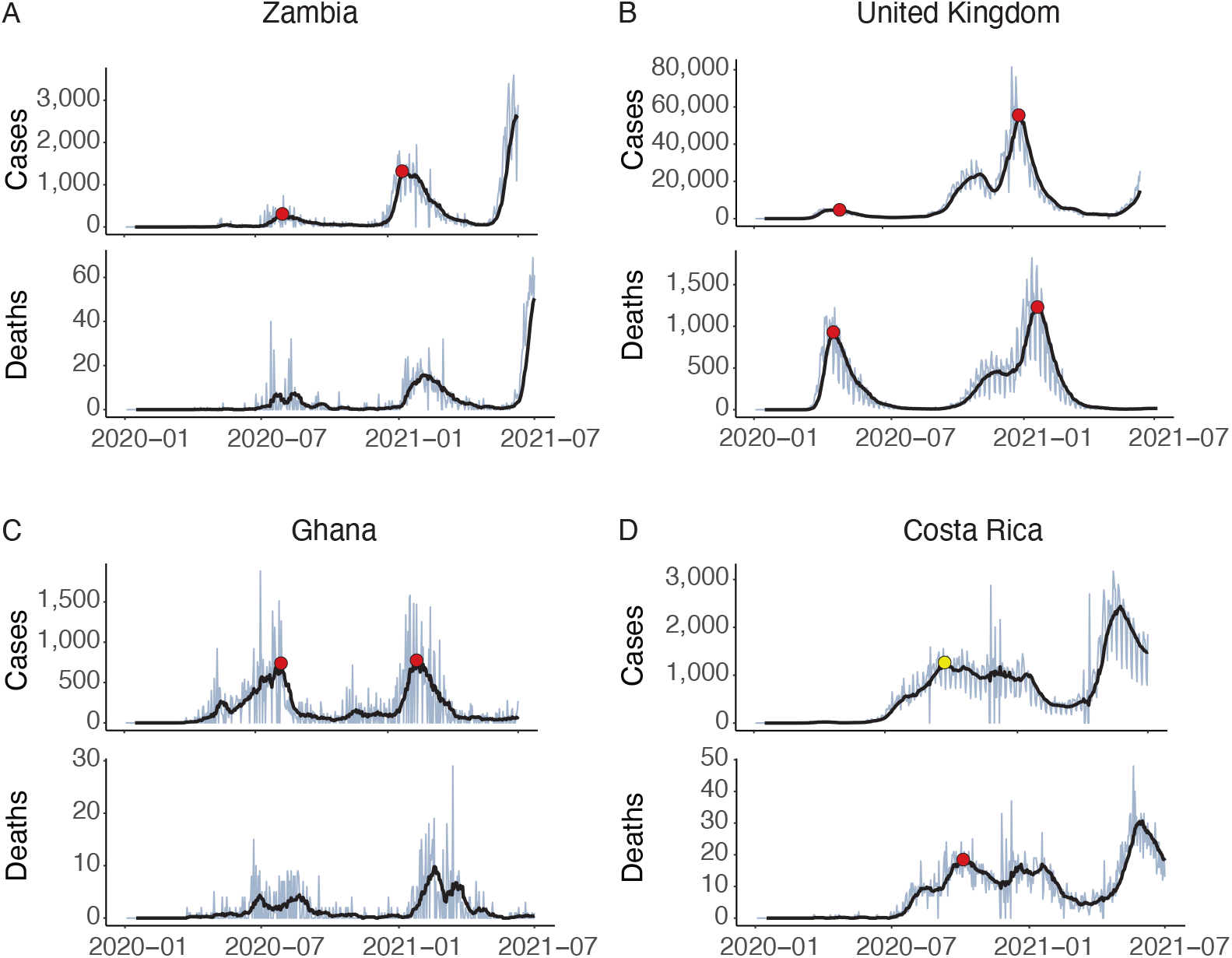
Identification of epidemic waves of COVID-19. **A:** Zambia shows a clear structure with two waves (red circles) in the cases data, while no waves are identified in the deaths data. **B:** the UK shows a structure that might arguably have two or three waves, but sub-algorithm D combines the final two. **C:** In Ghana sub-algorithm B filters out an early spike in cases. It is not clear visually whether this is noise or a meaningful epidemiological event; the algorithm cannot do better than the reader in determining this from simply inspecting a graph. No waves in deaths are identified due to low absolute counts. **D:** The number of cases in Costa Rica does not fall by 70% after the first wave, so it is not identified by the algorithm as a wave. This shows how important the parameter *P*_rel_ can be. However, cross-validating against the time series of deaths allows the wave to be identified (yellow circle)

There are a small number of outliers (Botswana, Cameroon, Equatorial Guinea, Republic of the Congo, and Western Sahara) for which a much larger number of waves are detected by the algorithm. On inspection, the time series for these countries are extremely noisy. This might be resolved by extending the smoothing window, but this would come at the cost of reduced accuracy for other countries. For this reason, we excluded these countries from the analysis. Countries with time series that never reached *P*_abs_ are also excluded from the analysis.

Table 2 reports certain epidemiological and socioeconomic characteristics according to the number of waves a country has experienced. Of the trends which can be observed there, only two are significant at the 5% level. A greater number of waves is associated with a longer stringency response time (a one-tailed Mann-Whitney test indicates that countries with more than one wave had significantly slower response time than those with only one wave, *p* = 0.0002) and with a higher GNI (*p* < 0.0001). The link to population density is not statistically significant.

**Table 2:** Basic characteristics of countries stratified by how many waves of COVID-19 they have experienced. The ‘Count’ variable indicates the number of countries in the cohort; not all characteristics are available for all countries in the cohort. The time t0 is the date of the tenth cumulative death.

|  | Countries with 1 wave |  | Countries with 2 waves |  | Countries with 3 or more waves |  |
| --- | --- | --- | --- | --- | --- | --- |
| Count | 57 |  | 84 |  | 51 |  |
|  | Median | IQR | Median | IQR | Median | IQR |
| Mortality (cumulative per 10,000) | 2.81 | 8.72 | 3.65 | 9.52 | 5.46 | 11.40 |
| Cumulative case rate (total confirmed per 10,000) | 196.99 | 609.05 | 233.18 | 698.56 | 353.62 | 675.67 |
| Peak case rate (peak new cases per day per 10,000) | 2.52 | 6.43 | 2.02 | 5.07 | 2.87 | 6.51 |
| Stringency response time (days from t0 to cancellation of public events) | -53.50 | 255.00 | -33.50 | 51.25 | -20.00 | 27.75 |
| Total stringency (integral under stringency index curve) | 26,601 | 6,867 | 30,062 | 8,186 | 31,413 | 5,6498 |
| Testing response time (days from t0 to 1 test per 1,000 cumulative) | 5.00 | 64.50 | 34.00 | 46.75 | 14.00 | 34.50 |
| Population density (people per square km) | 84.63 | 263.55 | 82.35 | 98.63 | 105.32 | 188.61 |
| GNI per capita (PPP adj 2017 US\$) | 12,191 | 17,806 | 13,459 | 24,079 | 20,083 | 34,832 |

Public discussion of waves of COVID-19 is common, with epidemiologists acknowledging the ambiguities around the term. Marc Lipsitch of Harvard has described a wave as “a useful metaphor” rather than a precise term, distinguishing waves from ‘momentary’ spikes [33] and Stephen Morse of Columbia has highlighted how epidemic waves in the traditional sense are “biological” phenomena, ending due to reduction in the susceptible population, while COVID-19 waves so far have ended due to measures “artificially designed to slow it down” [32].

This is not the first paper to seriously investigate the concept of observed epidemic waves. Taubenberger and Morens [28] note the three ‘explosive pandemic waves’ of 1918 influenza as a particularly unusual aspect of the pandemic. In the current context of COVID-19, seeking a rigorous means of identifying waves of COVID-19, Zhang et al. [35] propose an identification method which locates sustained periods of time when Rt is significantly larger than or less than 1. Values for Rt are obtained from the publicly available website [2] which uses a Kalman filter to estimate growth rate. Our method attends less to rates of increase and decrease as the defining aspect of a wave, focussing instead on identifying periods of high case numbers.

The shifts in sampling effort and sampling framework over the course of the COVID-19 pandemic greatly complicate the task of estimating the true level of infection. This paper contributes a semi-automated means of decomposing epidemiological time series into shorter periods, the waves. Without an algorithm of this type it is impossible to carry out the analysis above of how characteristics differ depending on the number of waves experienced. We also expect that analysis by wave period can reduce the impact of these shifts and provide customised time windows for each country studied. The method tallies well with natural intuition and allows for cross-validation between time series which the user expects to be correlated (cases and deaths). The parameters can be set by the user according to biological knowledge of the disease and the user’s own determination of how a wave should be defined in a particular context.

This method is grounded in a translation of visual intuition into mathematical rules and so it cannot perform better than a visual inspection of the time series. However, it does provide the advantage of setting an objective framework for identification of waves and the ability to process a large number of time series swiftly and consistently. Another aspect of this method is that every date in the time series is allocated to a wave, meaning every case and every death is allocated to a particular wave. This may not always be desirable. If there is a clear plateau between two waves then the location of the minimum which marks the end of one wave and the beginning of its successor will not be robust. A threshold test could easily be implemented to identify gaps between waves.

### 3.2 Wave typology

The typology of waves described in Table 3 describes the circumstances which may generate a time-series wave. Critically, these waves are not always driven simply by an increase in transmission.

**Table 3:**
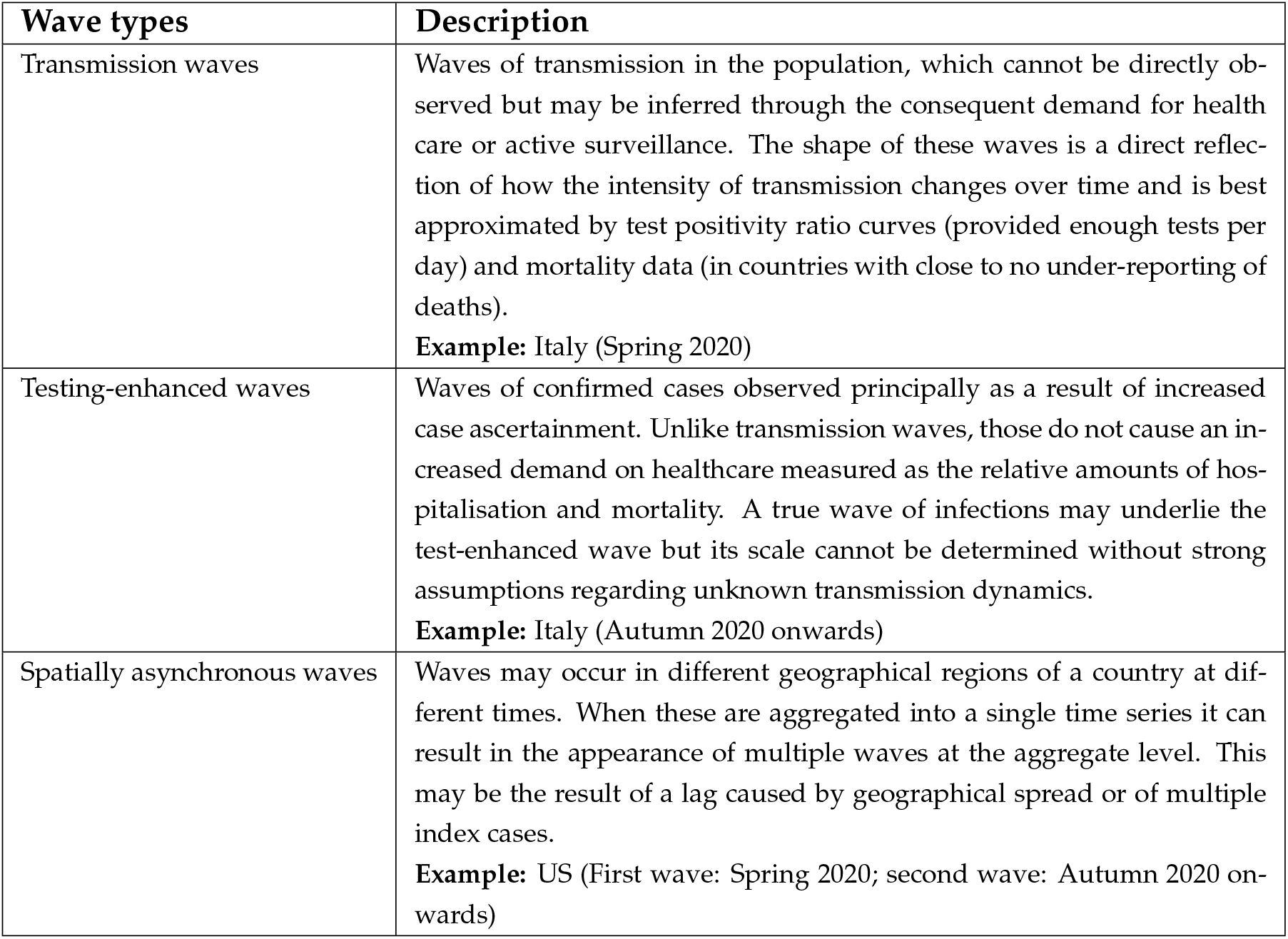
Types of waves of confirmed cases observed during the COVID-19 pandemic throughout 2020.

| Wave types | Description |
| --- | --- |
| Transmission waves | Waves of transmission in the population, which cannot be directly observed but may be inferred through the consequent demand for health care or active surveillance. The shape of these waves is a direct reflection of how the intensity of transmission changes over time and is best approximated by test positivity ratio curves (provided enough tests per day) and mortality data (in countries with close to no under-reporting of deaths).<br>**Example:** Italy (Spring 2020) |
| Testing-enhanced waves | Waves of confirmed cases observed principally as a result of increased case ascertainment. Unlike transmission waves, those do not cause an increased demand on healthcare measured as the relative amounts of hospitalisation and mortality. A true wave of infections may underlie the test-enhanced wave but its scale cannot be determined without strong assumptions regarding unknown transmission dynamics.<br>**Example:** Italy (Autumn 2020 onwards) |
| Spatially asynchronous waves | Waves may occur in different geographical regions of a country at different times. When these are aggregated into a single time series it can result in the appearance of multiple waves at the aggregate level. This may be the result of a lag caused by geographical spread or of multiple index cases.<br>**Example:** US (First wave: Spring 2020; second wave: Autumn 2020 onwards) |

These types were identified from direct comparison with the time series of deaths and of positivity rates. These descriptions are rooted in an assumption that the time series of deaths provides a more reliable and consistent indicator of trends in viral activity than the time series of cases. The two main drivers of waves in the case incidence times series are transmission and testing. A wave may be driven by an increase in transmission, an increase in testing, or, indeed, by a combination of these two drivers if the testing regime changes during a transmission wave. As a result, case incidence data from two subsequent waves can often not be usefully compared. The relative difference in drivers can be inferred, at least after the fact, from the presence or absence of an accompanying mortality incidence peak. We further identify a third type of wave at the national level (spatially asynchronous waves). Countries with this wave typology might benefit from evaluating the local epidemic curves in isolation and designing local intervention policies.

Figure 5 shows the structure of the waves of the epidemic for two example countries: Italy and the United States. In Italy there are two waves of confirmed cases and two waves of mortality, which occur at closely matched periods in time. However, the relative number of cases to deaths about each peak differs greatly from the first to the second wave, translating into a decreasing CFR (case fatality ratio) trend which needs to be critically engaged with. In the United States, we visually perceive three waves of cases and deaths, with the algorithm combining the first two wave structures into one wave. Again, there is a clear difference in the relative number of cases to deaths. In this particular instance, we observe geographical variation between the waves, with the outbreak being concentrated in different regions at different times (Figure 6). This is an example of spatially asynchronous waves.

**Figure 5:**
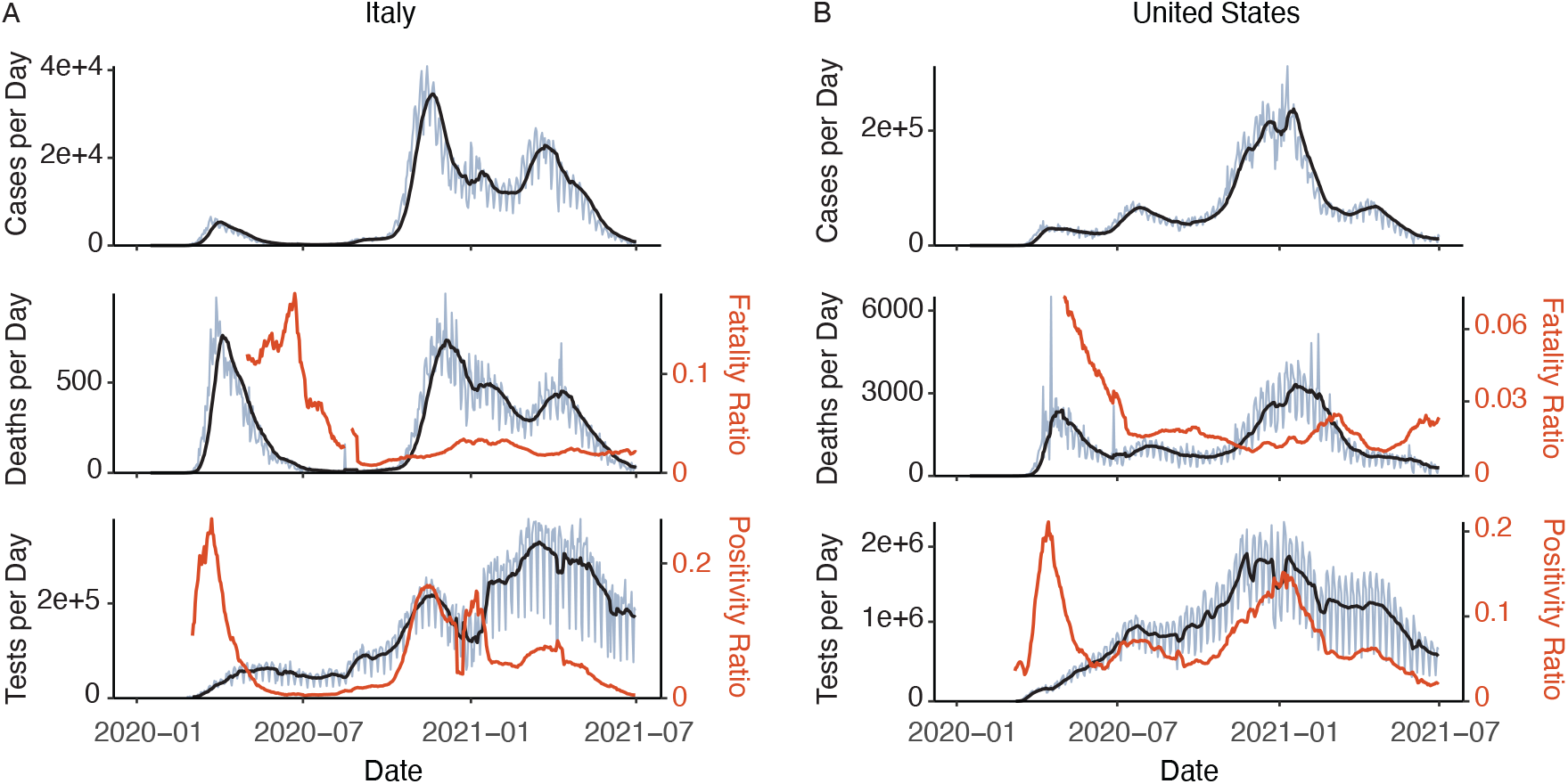
The structure of the waves of the epidemic for two example countries: Italy and the United States. In Italy there are two waves of confirmed cases and two waves of mortality, which occur at closely matched periods in time. However, the relative number of cases to deaths about each peak differs greatly from the first to the second wave, translating into a decreasing CFR (case fatality ratio) trend which needs to be critically engaged with. In the United States, we visually perceive three waves of cases and deaths, with the algorithm combining the first two wave structures into one wave. Again, there is a clear difference in the relative number of cases to deaths. In this particular instance, we observe geographical variation between the waves, with the outbreak being concentrated in different regions at different times. This is an example of spatially asynchronous waves. Daily counts are shown in light blue, with a 7-day average in black. Identified wave durations for daily cases are shown in green.

**Figure 6:**
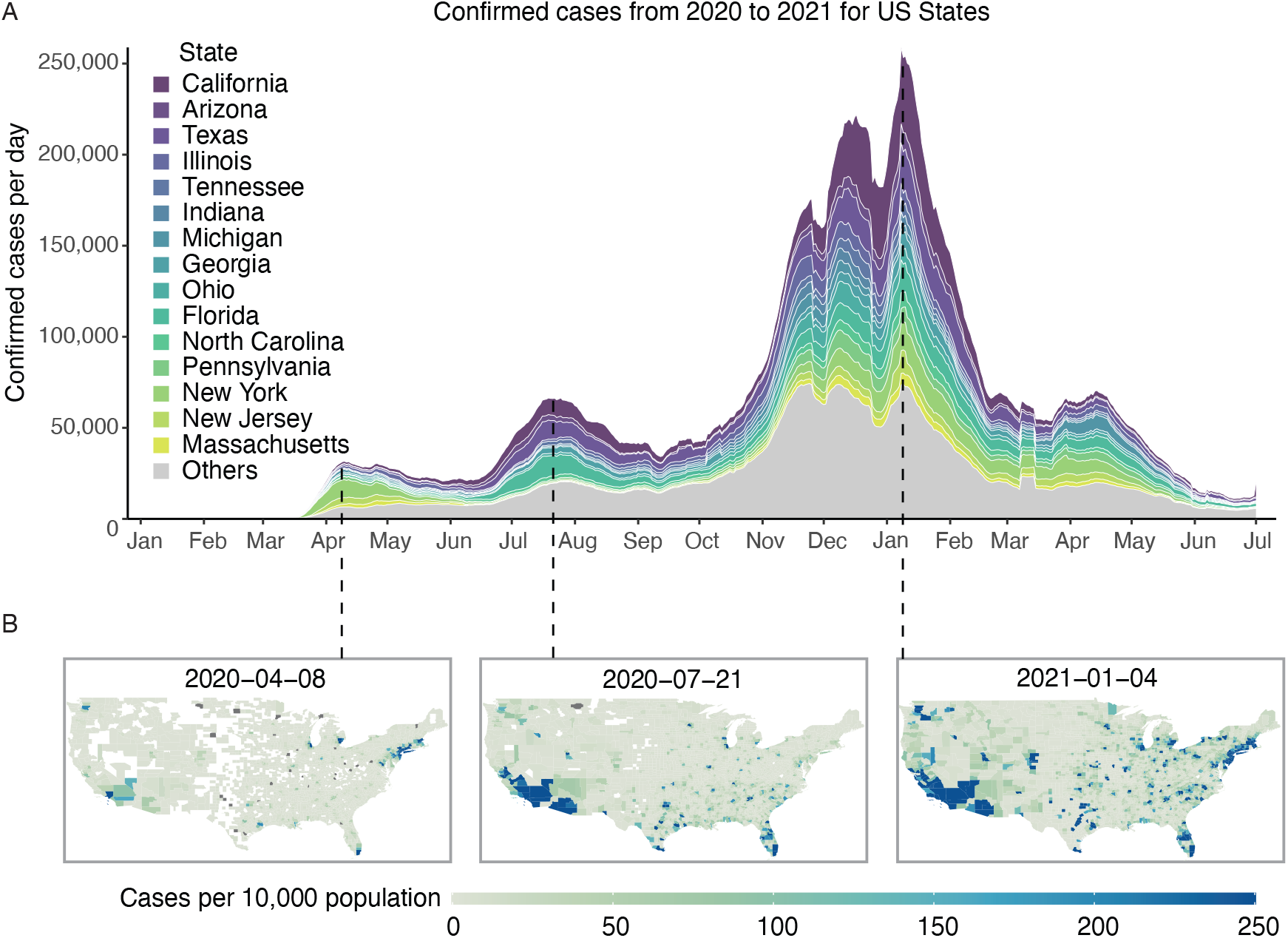
A visualization of how the geographical concentration of COVID-19 has varied over time across the United States. Individual states show wave structures but the result of aggregating data at the national level is that local waves overlap and are not so distinct from each other.

It is well known that as the criteria and/or capacity for testing for infection changes, the proportion of infections detected changes accordingly. Had a later version of the case definition for COVID-19 issued by the National Health Commission in China been in place from the start of the outbreak, 232,000 confirmed cases might have been reported in China by Feb 20, 2020, as opposed to the 55,508 cases actually reported, a 4-fold increase [29]. Also, most countries have vastly increased their testing capacity over time, so that the case ascertainment rate is now far greater than that in place when restrictions were initially introduced [14]. This creates difficulty in deciding how serious the present situation is and how it compares with the first wave of transmission as well as in describing the spread of the epidemic to the public [20] and has inspired researchers to propose methods of using mortality data as a more reliable proxy for infection numbers [9, 12, 18].

We find that the concept of spatially asynchronous waves is most relevant in large countries. However, even in a small country, the progression of case numbers can usefully be thought of as the result of several superimposed outbreaks separated in both time and space, creating wave patterns in case numbers which may not fit our definition of a wave. [19] found that a classification of countries according to the number of subepidemics and whether their intensity increases or decreases was well explained by government action, with declining subepidemics associated with stringent action and increasing subepidemics associated with less stringent action.

### 3.3 Modulation of waves by NPIs

The stringency and speed of the introduction of NPIs and large-scale testing were compared to the severity of epidemic waves (as measured by the per capita death rate during that wave).

To account for differences in epidemic start date, measures of time are calculated relative to the date at which each country first reached 10 cumulative deaths (referred to in this paper as start date T0). Reducing this threshold would give undue weight to importation events at the expense of locally acquired infections. Smaller countries (population ¡ 2.5 million) were excluded from the analysis as T0 was often very late in these countries.

Table 4 shows the relationship between the features of a government’s response and the per capita death rate in that country. Table 5 shows the relationship between testing levels and the per capita death rate in that country. Both tables show positive correlations between the mortality burden and the overall scale of interventions or testing, which are strengthened when the relationships are studied by waves, an analysis made possible by the wave identification algorithm. We must urge caution in the interpretation of this observed correlation. The feedback process between the use of NPIs and mortality is complex and we have not carried out a causal analysis of this relationship. Most likely, our results suggest that countries that have introduced more stringent and/or prolonged measures did so in response to higher mortality figures, not that higher stringency measures caused more deaths.

**Table 4:** Kendall’s rank correlation between the mortality burden of the pandemic in different waves and aspects of government response. Stringency Response Time is defined to be the time from T0 until public events were cancelled. The analysis is carried out for alternative definitions of Stringency Response Time in the Supplementary Material. Total Stringency and Wave Stringency are the integrals of the stringency index over the relevant period. Epidemic phase is an ordinal variable indicating which wave the country is currently in, and whether it has passed the peak of its current wave. The analyses for Wave 2 Deaths were carried out only for countries which experienced a second wave.

| Burden metric | Stringency metric | Kendall's rank correlation | p-value |
| --- | --- | --- | --- |
| Wave 1 Deaths | Stringency Response Time | 0.24 | <0.0001 |
| Wave 2 Deaths | Stringency Response Time | 0.23 | 0.0019 |
| All Deaths | Stringency Response Time | 0.34 | <0.0001 |
| Epidemic Phase | Stringency Response Time | 0.24 | 0.0003 |
| Wave 1 Deaths | Total Stringency | 0.25 | <0.0001 |
| Wave 2 Deaths | Total Stringency | 0.13 | 0.082 |
| All Deaths | Total Stringency | 0.20 | 0.0005 |
| Wave 1 Deaths | Wave 1 Stringency | 0.32 | <0.0001 |
| Wave 2 Deaths | Wave 2 Stringency | 0.38 | <0.0001 |

**Table 5:** Kendall’s rank correlation between the mortality burden of the pandemic in different waves and aspects of testing response. Testing Response Time is the time from T0 until reaching a threshold for the level of testing (1 test per 1000 population). The analysis is carried out for alternative definitions of Testing Response Time in the Supplementary Material. Epidemic phase is an ordinal variable indicating which wave the country is currently in, and whether it has passed the peak of its current wave. The analyses for Wave 2 Deaths were carried out only for countries which experienced a second wave

| Burden metric | Testing metric | Kendall's rank correlation | p-value |
| --- | --- | --- | --- |
| Wave 1 Deaths | Testing Response Time | 0.18 | 0.013 |
| Wave 2 Deaths | Testing Response Time | -0.022 | 0.79 |
| All Deaths | Testing Response Time | 0.14 | 0.047 |
| Epidemic Phase | Testing Response Time | 0.021 | 0.78 |
| Wave 1 Deaths | Total Tests | 0.26 | 0.0005 |
| Wave 2 Deaths | Total Tests | 0.41 | <0.0001 |
| All Deaths | Total Tests | 0.42 | <0.0001 |
| Wave 1 Deaths | Wave 1 Tests | 0.42 | <0.0001 |
| Wave 2 Deaths | Wave 2 Tests | 0.37 | 0.0033 |

There is also a positive correlation between the time taken to implement the interventions or testing and the mortality burden. In the case of NPIs, this correlation is stronger for the overall burden than it is for the individual waves, which can be explained by the positive correlation between the response time and the number of waves experienced. The time to implement these measures might not be expected to influence the burden of the second wave of an epidemic, since it is distant in time. However, we do find that the correlation between time to implement NPIs and the burden of the second wave is similar to that for the burden of the first wave. This may be because response time influenced conditions at the beginning of the second wave, or it may indicate a country’s general policy approach. No significant relationship between testing response time and the burden of the second wave was found.

Although some early NPIs such as implementation of testing strategies, isolation of confirmed cases and tracing of their contacts slowed down early transmission, in most places these measures were not sufficient to contain the outbreak [8, 16]. Studies have found that while moderate measures are capable of reducing the size of the ensuing epidemic, severe measures are needed to suppress an epidemic so as to avoid overwhelming healthcare systems [7].

The positive correlation between the strength of interventions and the burden of the pandemic has been observed before. This relationship has been the subject of lively debate centered on the difficulty in collecting reliable data, the importance of timing, and the fact that interventions are an endogenous variable, with governments implementing interventions in response to current or anticipated case load [3]. Indeed, attending more closely to the timing of interventions and effects on the transmission rate rather than on absolute case numbers demonstrates that a wide range of restrictions are associated with a reduction in *Rt* [4, 17, 15], with early implementation being crucial.

Our findings provide further confirmation of the importance of early introduction of NPIs. Countries which introduced early NPIs tended to experience a lower mortality burden in the first wave, were less likely to experience a second wave and, if a second wave was experienced, that wave came with a lower mortality burden. These three effects combine to produce a very strong relationship between early introduction and low overall mortality.

## 4 Conclusion

It is possible to distill the natural visual understanding of ‘waves’ in a time series into simple mathematical rules which can be used to objectively annotate a large number of time series, identifying their component waves. In the context of COVID-19, these waves may result from increased transmission, increased testing, or some combination of the two. Not only this, waves can occur due to the aggregation of time series from a large geographical area, so that the second wave is really a first wave, but for a different region of the country. When engaging in comparative study of the relationships between interventions taken and the mortality burden of the disease, using the wave as the time unit for analysis can result in clearer conclusions. The speed with which interventions are implemented is strongly correlated with the wave structure of the subsequent epidemic.

## Data Availability

All data produced are available online at
https://github.com/covid19db
https://ourworldindata.org/

